# Using functional near-infrared spectroscopy to explore neurocognitive function in adult survivors of childhood acute lymphoblastic leukemia

**DOI:** 10.1101/2024.09.03.24312978

**Authors:** Simon Skau, Marianne Jarfelt, Gustaf Glavå, Laura Jess, H. Georg Kuhn

## Abstract

Acute lymphoblastic leukemia (ALL) is the most common childhood malignancy. Due to the drastic increase in survivor rate over the last 50 years, long lasting treatment effect on moods and neurocognitive function has become a present issue. Most studies of late effects of treatment of ALL survivors investigate patients in their adolescents. This pilot study aims to identify measurements for evaluating late effect of childhood ALL survivors regarding neurocognitive and mood problems in adulthood. ALL survivors who received neurotoxic treatment with high-dose methotrexate and cranial radiotherapy (Chemo+CRT) (n=10) and ALL survivors only treated with high-dose methotrexate (Chemo) (n=10), plus age and sex match controls (n=20) where recruited to the study. The study protocol involved questionnaires, neurocognitive tests and optical brain imaging with functional near infrared spectroscopy (fNIRS) over the frontal and parietal cortex. The fNIRS results indicate a reduced involvement of the parietal cortex during conflict processing for the ALL survivors compared to controls. The study protocol shows promising results for identifying subgroups that suffers from neurocognitive and mood problems and we aim to expand upon it in a larger study. As our results indicate increased challenges among female ALL survivors, especially pathological fatigue, anxiety, and information processing, it is important to explore in future investigations the interplay between the risk of hormonal interaction with chemotherapy during development and occupational and social pressure during adulthood.

## 1. INTRODUCTION

Acute lymphoblastic leukemia (ALL) is the most common childhood malignancy, accounting for approximately 25-30% of pediatric malignancies[1]. The survival rate for pediatric ALL has improved from 5% to over 90% over the last 50 years in high income countries [2-4]. Cranial radiotherapy (CRT) improved the prognosis during the 1970 and 1980s but resulted in neurocognitive and endocrine complications, together with secondary malignancies [5, 6]. Therefore, high-dose methotrexate has replaced CRT in most protocols except for patients with primary CNS leukemia or relapse in the CNS. In the Nordic countries, the first treatment protocol to avoid CNS irradiation except for those with very high-risk disease or CNS involvement at diagnosis, was launched in 1992 [7]. However, with time it became apparent that neurocognitive and psychological complications are still present in this group [8, 9]. It has been estimated that 50% of individuals treated with high-dose methotrexate experience late neurocognitive problems [10], with an increased risk for female survivors [11].

Studies on neurocognitive functions in childhood ALL survivors have reported heterogeneous results. The problems identified during adolescence are lower IQ, executive function and processing speed compared to controls [10-14], but not necessarily against normalized populations [9]. Reported symptoms are in line with those of attention-deficit/hyperactivity disorder (ADHD) [15, 16]; however, cognitive testing has not been able to confirm problems with attention or concentration [12, 13] and mixed result were reported regarding processing speed [12]. Other symptoms experienced by childhood ALL survivors with neurocognitive problems during adolescence include sleep problems and fatigue [10], with females having a higher risk than males [17]. One previous study in long-term adult survivors of childhood ALL found that prevalence of fatigue was associated with higher age at study (> 30y) and older age at diagnosis (>6 y)[18]. Although this suggests an exacerbated fatigue risk in adulthood, most previous research about fatigue has studied children or adolescents relatively recent after completed ALL treatment [9, 10, 19].

Brain imaging studies have also been used to detect the neurocognitive problems experienced by childhood ALL survivors. In a recent literature review on late brain effects after chemotherapy alone, Gandy *et al*. identified both structural and functional problems [20]. Structurally, reduced grey matter volume in all four lobes was detected in conjunction with reduced white matter volume in the frontal, parietal and temporal lobes, cortical thinning in the frontal and medial parietal lobes, and with greater reductions in female cancer survivors [20]. Childhood ALL survivors who experience neurocognitive problems exhibit reduced functional connectivity in the default mode network and reduced functional activity during sustained attention tasks in parietal and temporal cortex [20]. Of the 23 articles reviewed, 21 studies included participants with a mean age between 10 and 14 years. The two additional studies with longer assessment times (mean age 23-25 years) found reduced white matter in the frontal and temporal lobes and that patients treated with dexamethasone had reduced functional activity in the retrosplenial brain region [21, 22].

In previous studies our protocols have identified functional and cognitive problems related to pathological fatigue in adults that suffers from traumatic brain injury and exhaustion disorder [23, 24]. Pathological fatigue is characterized by the sensation of fatigue or disposition of fatigability interfering with common and desired activities such as working/going to school or social life [25]. The brain imaging method used in this study is functional near-infrared spectroscopy (fNIRS). fNIRS uses near-infrared light to measure the relative change in oxygenated and deoxygenated hemoglobin (oxy-Hb/deoxy-Hb) in the cortex as an indirect measure of neural activity [26, 27]. The aim of this pilot study was to determine whether a set of questionnaires, cognitive tests and functional brain imaging could identify subgroups of long-term adult childhood ALL survivors who suffer from neurocognitive impairments in order to help understand the underlying neurocognitive mechanism.

## 2. MATERIALS AND METHODS

### 2.1 Study participants and protocol

Adult childhood ALL survivors who had visited the long-term follow-up clinic at Sahlgrenska University hospital and who met the inclusion criteria were asked to participate. Patients were eligible if they were aged ≥ 18 years, diagnosed with ALL during childhood (< 18 years of age), and at least five years post-treatment at time of recruitment and had at least one visit to the Sahlgrenska long-term follow-up clinic prior to the study. Exclusion criteria: Ongoing recurrence or second malignancy, stem cell transplantation, impaired communicative ability, or insufficient knowledge of the Swedish language. All participants received written and oral information about the study and written consent was obtained prior to the investigation. Healthy controls where requited recruited in a convenience sampling. Exclusion criteria were a score below 26 on the Montreal Cognitive Assessment (MoCA) and inability to follow instruction. This study was approved by the Swedish ethics review authority (Etikprövningsmyndigheten – Dnr 2020-01580).

### 2.2 Experimental design

All participants performed the tasks in the same order (Figure 1A), starting with the following questionaries (VAS, MFS, and HAD), then pen and paper tasks: Montreal Cognitive Assessment (MoCA) [28], Symbol Search (SS) [29], Digit Symbol Coding (DSC) [29], followed by digit span forward and backwards. Thereafter, they performed a reaction time task (RT), the AX-Continues performance task (AX-CPT), followed by the Color Word matching Stroop task (CWM-Stroop test) and an on-screen spatial navigation task (not presented in this paper). Individual participants were seated in a chair at a table with a computer screen and executed all tests under same settings. An elastic cap with fNIRS optodes and detectors attached was carefully placed on the participant’s head. fNIRS recording was performed during the spatial navigation task and the CWM-Stroop test. The test procedure took between and 90-100 minutes, including preparatory work and practice trials for all tasks.

**Figure 1.**
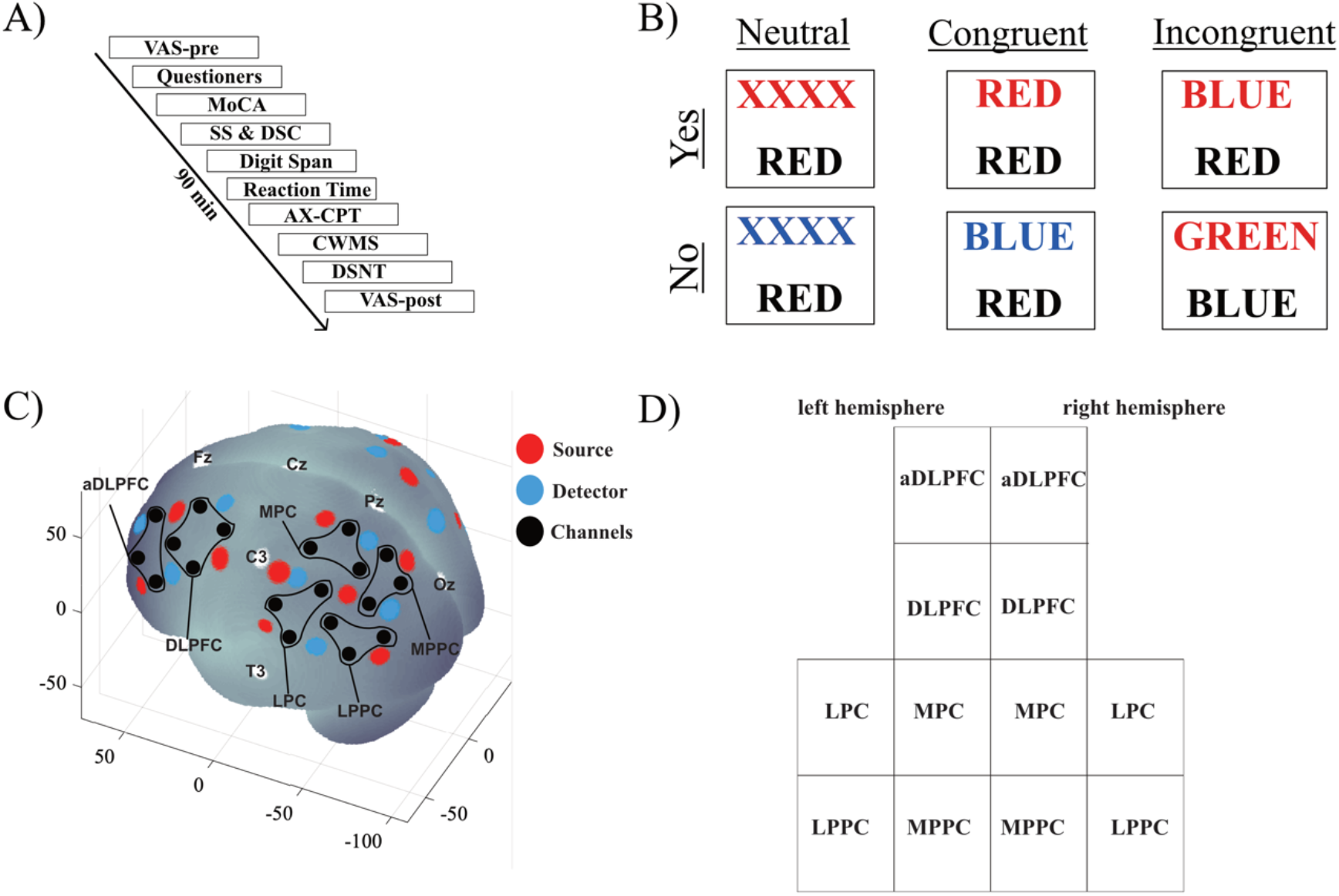
**A)** Experimental protocol. VAS – visual analog scale, MoCA – Montreal Cognitive Assessment, SS – Symbol Search, DSC – Digit Symbol Coding, AX-CPT – AX Continuous Performance Task, CWM-Stroop test – Color-word matching Stroop task, DSNT – Dresden Spatial Navigation Task. **B)** The different conditions during the CWM-Stroop test. **C)** Location of the fNIRS optodes. Red dots are light sources, blue dots are detectors, black dots are estimated channels, white dots are 10/20 landmarks. Black lines areas indicate the regions of interest (ROIs): MPPC – medial posterior parietal cortex or Brodmann area (BA) 7, LPPC – lateral posterior parietal cortex BA 7 and 40, MPC – medial parietal cortex BA 2 and 5, LPC – lateral parietal cortex, DLPFC – dorsolateral prefrontal cortex BA 8 and 9, aDLPFC – anterior dorsolateral prefrontal cortex BA 9 and 46. The axes refer to the MNI (Montreal Neurological Institute) coordinate system. The brain image was generated by a MATLAB-based toolbox[30]. **D)** Visual layout of the ROIs used in the data presentation (Figure 4) corresponding to the channel layout in **C)**.

### 2.3 Measurements

Before and after the experimental procedure, participants rated their state fatigue level on a visual analog scale (VAS). The VAS scale was a continuous line (10 cm) between the two end-points: “Pigg” (fresh/sharp) and “totally exhausted”.

Mental fatigue scale (MFS). The MFS is comprised of fourteen questions that are scored between 0 and 3 points, plus an additional question. The 42-point scale has a recommended cutoff value of 10.5 to indicate pathological MF [31, 32]. The questionnaire inquiries about fatigue in general, lack of initiative, mental recovery, concentration difficulties, memory problems, slowness of thought, sensitivity to stress, emotional instability, irritability, sensitivity to noise and light, and sleep. The MFS is unaffected by age, sex, or education [31, 33].

The Hospital Anxiety and Depression Scale (HAD) [34]. The HAD comprises fourteen questions with scores from 0 to 3 (scoring is not visible to the participants), seven questions each regarding trait anxiety and trait depression. Maximal score is 21 for each subscale, with a score of 7 indicating mild anxiety or mild depression, respectively and a score of 10 or higher is indicator of severe anxiety or depression, respectively.

To assess the participants’ global cognitive capacity, we used the Montreal Cognitive Assessment (MoCA). The MoCA is a fast screening instrument to detect mild cognitive impairment and dementia and confirm that the participants were cognitively healthy [28].

Attention, mental and psychomotor operation speed, and visual discrimination were measured using Symbol Search (SS) and Digit Symbol Coding (DSC). SS and DSC are subtests within the Processing Speed Index of the Wechsler Adult Intelligence Scale WAIS-IV [29]. In both tests, the subject was asked to correctly identify and decode as many symbols as possible for 2 minutes. Based on the two tests, a Processing Speed Index (PSI) is calculated using age-normative values.

Digit Span was used to measure working memory. The researcher read a sting of digits and the participant had to report the digits in the same order (for the forward task) or in reverse order (for the backward task). If the participants succeeded in a trial, then the next trial expanded the sequence by one digit). The participants had a maximum of four trials one each level. The task ended either after responding incorrectly on all four occasions at a span length or after successfully responding to 9 digits in a row. Digit Span raw score is defined as the length of the last successful digit span. The average of the raw scores from Digit span forward and backward are used in the analysis.

Reaction time. The participants were asked to press the space key as fast as possible on a computer keyboard whenever they heard a sound with a 5-second interval.

AX Continuous Performance Task (AX-CPT) was used to measure reactive and proactive cognitive control [35]. The participants were presented with a series of individual letters in the center of a computer screen from the following set of letters: A, B, D, G, H, J, K, V, X, Y, Z. the trials are divided in to cues and target items. The cue item informs the participants how they should respond to the target item. There are two cue items called A and B (non-A), and two target item called X and Y (non-X). The participant was instructed to respond by pressing a button with the left hand whenever an “X” target was preceded by an “A” cue, and answer with their right hand for all other trials. This cue-target pair is hereafter referred to as probe. The AX pair is called a “target probe”. For all other probes, BX (any non-“A” followed by an “X”), BY (any non-“A” followed by a “non-X” and AY(“A” followed by any non-“X”), a response with the right hand was required [36]. This generated four different probe types: AX (target), AY (reactive control beneficiary situation), BX (proactive control beneficiary situation) and BY (control). There were 150 probes with a distribution of 70% AX probes, 10% AY probes, 10% BX probes and 10% BY probes. There were 3 seconds between each cue and target trial, and 1 second between each target and cue trial. In total there were 298 trials, and a 30 second pause was given after half of the trials. Prior to the test, participants performed a practice run of the task with 50 trials. Raw scores recorded were reaction time, errors, and omissions.

From the response time data, we calculated the proactive behavioral index (PBI) according to the formula below, which had previously been applied in similar contexts [35, 37, 38]:

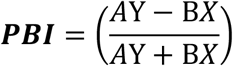

A positive value indicates a more proactive tendency and a negative value a more reactive tendency of problem solving.

Color-word matching Stroop task (CWM-Stroop test) [39]. In the CWM-Stroop test participants are presented with two rows of letters/words on a computer screen. The participants are instructed to determine whether the ink color of the letters in the top row matches the color name written in the lower row (Figure 1B). Answers were given by pressing a button on a gamepad, with a press with the right hand for Yes and with the left for No. The words on the lower row were always printed in black, and the color name used where RÖD, GRÖN, GUL and BLÅ (Swedish words for red, green, yellow and blue respectably). There were three types of trials. For neutral trials, the letters in the top row were XXXX printed in red, green, blue or yellow ink. For congruent and incongruent trials, the top row consisted of the words RED, GREEN, YELLOW, and BLUE, and printed in the matching color for the congruent trial and printed a non-matching color for the incongruent trial. To shift visual attention to the word in the top row, it was presented 350 milliseconds before the lower word.

In total there where 60 trials (20 neutral, 20 congruent, and 20 incongruent) trials with a 12-second interstimulus interval. Each condition had an equal number of Yes and No answers. Words remained on the screen until the response was given within a maximum time of 3 sec. A “+” symbol was shown between trials.

### 2.4 fNIRS data acquisition

The fNIRS measurements were performed using a continuous wave system (NTS, Optical Imaging System, Gowerlabs Ltd., UK) [40] with two wavelengths (780 and 850nm) in order to measure changes in the concentration of oxygenated hemoglobin (oxy-Hb) and deoxygenated hemoglobin (deoxy-Hb). The system has 16 dual-wavelength sources and 16 detectors. The array used 38 channels (i.e., source/detector pairs) with a source-detector distance of 30 mm and two short-separation channels with a 10-mm distance, as suggested by previous studies [41, 42]. Short separation channels are only sensitive to hemodynamic activity in the scalp and skull. Since the regular channels measure signals originating in the brain and the scalp and skull, the use of short-separation channels allows to regress out the scalp signal to improve the specificity of the fNIRS measurement for hemodynamic responses from within the brain [41, 42]. The optode placements were designed to encompass both the left and right DLPFC and the parietal cortex (Figure 1C). The fNIRS data were acquired at a sampling frequency of 10 Hz.

### 2.5 fNIRS data analysis

The fNIRS data were preprocessed using MATLAB 2018b [43] and the MATLAB-based fNIRS-processing package Homer3 [44]. Trials with response errors or omission CWM-Stroop test were eliminated before preprocessing. The processing pipeline started with pruning channels usingHomER3 function *hmrR_PruneChannels*, where channel were rejected if their mean intensity was below the instrument’s noise floor (1e-3 AU) and the standard deviation of the a signal to noise ratio was above 15. Raw data were then converted to optical densities. A high band-pass filter of 0.05 Hz was used to correct for drift and a low band-pass 0.5 Hz filter to remove pulse and respiration. Examples of the hemodynamic response curves are visualized in Supplementary Figure 1.

The functions *hmrR_MotionArtifact* and *hmrR_MotionCorrectSpline* with the recommended value of 0.99 were used to correct for motion artifacts. The function *hmrOD2Conc* was used to convert optical density to hemoglobin concentration with partial pathlength factors for each wavelength set according to age based on [45]. To calculate the hemodynamic response function (HRF), the *hmrR_GLM* function in HomER3 was used, which estimates the HRF by applying a General Linear Model (GLM). To solve the GLM, an iterative weighted least-square fit of a convolution model in which the HRF at each channel and chromophore was modeled as a series of Gaussian basis functions, with a spacing and standard deviation of 0.5 seconds [46]. The model did not include any polynomial drift regressors due to the use of a high bandpass filter. The regression time length was –2 to 12 seconds. Channels were then averaged across predefined regions of interest (ROIs) (see Figure 1C).

### 2.6 Statistics

We used MATLAB 2018b [43] and the open-source program *JASP* version 0.16.4 [47] for statistical analysis and visualization. Since we consider this study a pilot and exploratory study, no generalizing statistic is presented, instead for behavioral and demographic data, summary statistics such as mean and standard deviation are used.

For fNIRS data, the maximum peak between 3 and 9 seconds after each stimulus was identified for each ROI, and the two seconds around the peak value were averaged. Comparison is done with independent t-test, and only t-values are presented.

In Figures 2-4 we use 95% confidence intervals (CI) as error bars. Although the CI could be used to generalization to a larger population, no such attempt is intended due to the exploratory nature of the study. Summary statistic of all data is presented in the Supplementary materials.

**Figure 2.**
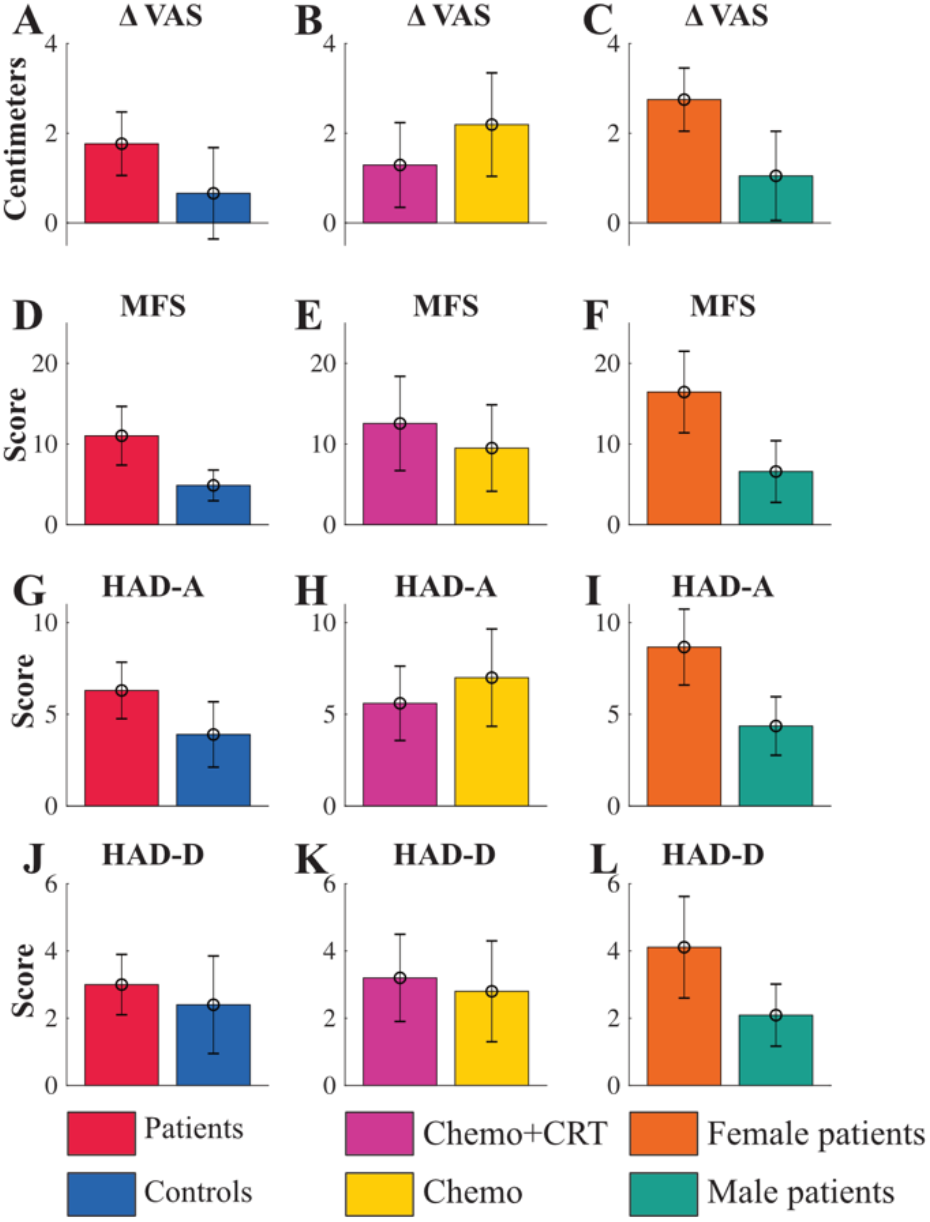
**A-C** show delta VAS (visual analogue scale) for Controls and ALL survivors (A), Chemo+CRT and Chemo (B) and female and male ALL survivors (C). D**-F** show MFS (mental fatigue scale) for Controls and ALL survivors (D), Chemo+CRT and Chemo (E) and female and male ALL survivors (F). **G-I** show anxiety scale from the HAD (Hospital Anxiety and Depression) scale for Controls and ALL survivors (G), Chemo+CRT and Chemo (H) and female and male ALL survivors (I). J**-L** show depression scale from the HAD scale for Controls and ALL survivors (J), Chemo+CRT and Chemo (K) and female and male ALL survivors (L). Error bars are confidence intervals.

## 3. RESULTS

### 3.1 Participants

A total of 25 adult childhood ALL survivors were invited of whom 20 participated (10 treated with CRT and 10 without CRT). Treatment data of the participating ALL survivors are presented in Table 1. Among those who did not participate, three had been treated with CRT and 2 had not. Demographical and clinical characteristics of the study population are presented in Table 2. The controls were recruited with a convenience sampling method from hospital staff and friends. Despite the use of convenience sampling, there were no significant differences in age, education level, nicotine and alcohol use, or previous psychiatric problems between the ALL-survivor groups and controls (see Table 2). The only difference between groups was the number of individuals engaged in full-time employment. In the Chemo+ CRT group, only 6 out of 10 individuals worked full-time, while all individuals in the Chemo group and 19 out of 20 individuals in the control group worked full-time.

**Table 1.** Treatment data of the participating ALL survivors.

|  |  | Mean | SD | Min | Max |
| --- | --- | --- | --- | --- | --- |
| Age at diagnosis | Chemo+CRT | 8.0 | 4.5 | 2.0 | 16.0 |
|  | Chemo | 7.5 | 5.0 | 2.0 | 16.0 |
| Year since end of diagnosis | Chemo+CRT | 23.5 | 6.3 | 15.0 | 32.0 |
|  | Chemo | 21.1 | 5.7 | 6.0 | 27.0 |
| Radiation dose (Gy) | Chemo+CRT | 19.0 | 3.16 | 18.0 | 28.0 |
|  | Chemo | - | - | - | - |
| Mtx dose iv (mg/m<sup>2</sup>) | Chemo+CRT | 8530 | 14841 | 0 | 48300 |
|  | Chemo | 36500 | 6851 | 25000 | 45000 |
| Mtx dose it (mg) | Chemo+CRT | 125 | 90 | 36 | 288 |
|  | Chemo | 168 | 34 | 132 | 252 |
| Cyclofosfamide dose iv (mg/m<sup>2</sup>) | Chemo+CRT | 2540 | 2162 | 0 | 6600 |
|  | Chemo | 2300 | 1252 | 0 | 3000 |
| Prednisolone (mg/m<sup>2</sup>) | Chemo+CRT | 3988 | 922 | 1680 | 5220 |
|  | Chemo | 2412 | 1122 | 1680 | 4620 |
| Dexamethasone (mg/m<sup>2</sup>) | Chemo+CRT | 201 | 119 | 0 | 400 |
|  | Chemo | 316 | 229 | 0 | 800 |

Cumulative radiation dose expressed as Gray (Gy). Cumulative dose of chemotherapy expressed as milligram per meter square (mg/m<sup>2</sup>) for intravenous (iv) chemotherapy and oral corticosteroids and in milligram (mg) for intrathecal (it) methotrexate. SD, standard deviation; Min, minimum value; Max, Maximum value.

**Table 2.**
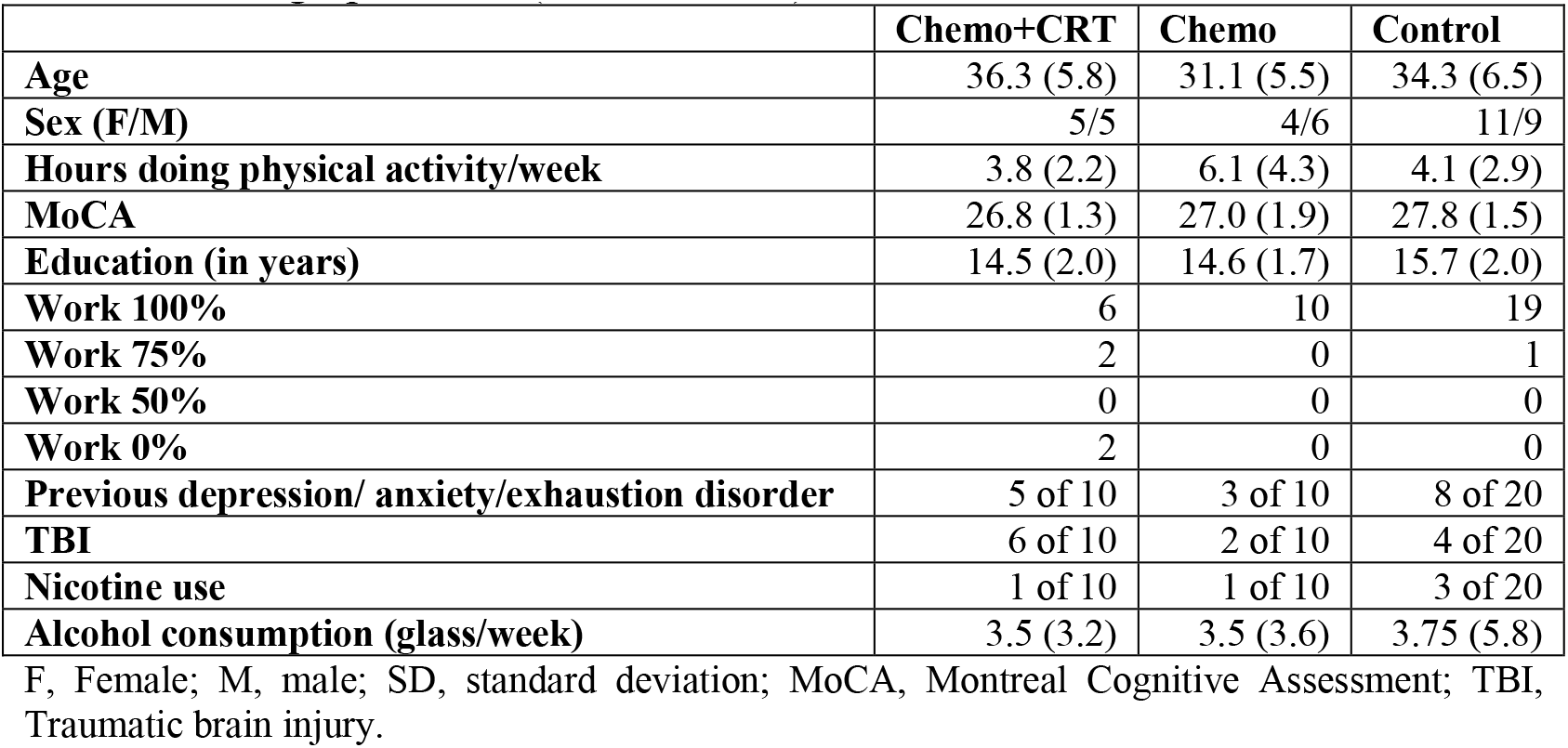
Demographic data (mean and SD)

### 3.2 Questionnaires

Overall, ALL survivors reported a greater change in state fatigue during the cognitive assessment session compared to controls as measured by delta VAS (Figure 2A). When comparing the treatment groups, the Chemo group showed a higher delta VAS than the Chemo+CRT group (Figure 2B) due to the fact that the Chemo+CRT group reported a lower state fatigue already at the start of the test session. Chemo-CRT, Chemo and Controls pre-VAS were pre 3.8±3.4, 2.6±1.9 and 2.6±1.6, and their post-VAS were 5.4±3, 4.8 ±2.3 and 3.1±2.0, respectively. As illustrated in the Figure 2C, females got more fatigued by the procedures in both ALL survivor groups.

The ALL survivors reported higher trait fatigue scores on the MFS than controls, but with a large heterogeneity (Figure 2D), since the Chemo+CRT showed higher trait fatigue levels than the Chemo group (Figure 2E). The largest difference was observed between the sexes (Figure 2F), with seven out of nine female ALL survivors reporting fatigue levels above the recommended cutoff (10.5 as an indication of pathological fatigue), while only one out of eleven male ALL survivors did.

Similarly, ALL survivors reported a higher level of trait anxiety compared to controls, as detected by the HAD-A questionnaire (Figure 2G). The Chemo group reported a slightly higher anxiety level than the Chemo+CRT group (Figure 2H). A sex difference was also present with seven out of nine women reporting anxiety levels above cutoff of 7 (7 indicator of mild anxiety problem, 10 severe anxiety problems), whereas only two out of eleven men scored above 7 (Figure 2I). For depressive symptoms there were no group difference, neither between ALL survivors and controls, between Chemo+CRT and Chemo, nor between men and women (Figure 2J-L). None of the groups reported problems with depression (score above 7), except for two individuals in the control group.

### 3.3 Behavioral measurements

There were no differences in a simple reaction time task or in digit span, with rather homogeneous result between all subgroups (Figure 3A-F). For the Processing Speed Index (PSI), there was no overall difference between patients and controls (Figure 3G). On average, the Chemo group scored close to the 75th percentile and the controls scored above the 63rd percentiles for their age. The Chemo+CRT group performed on average in the 25th percentile for their age which was worse than the Chemo group (Figure 3H).

**Figure 3.**
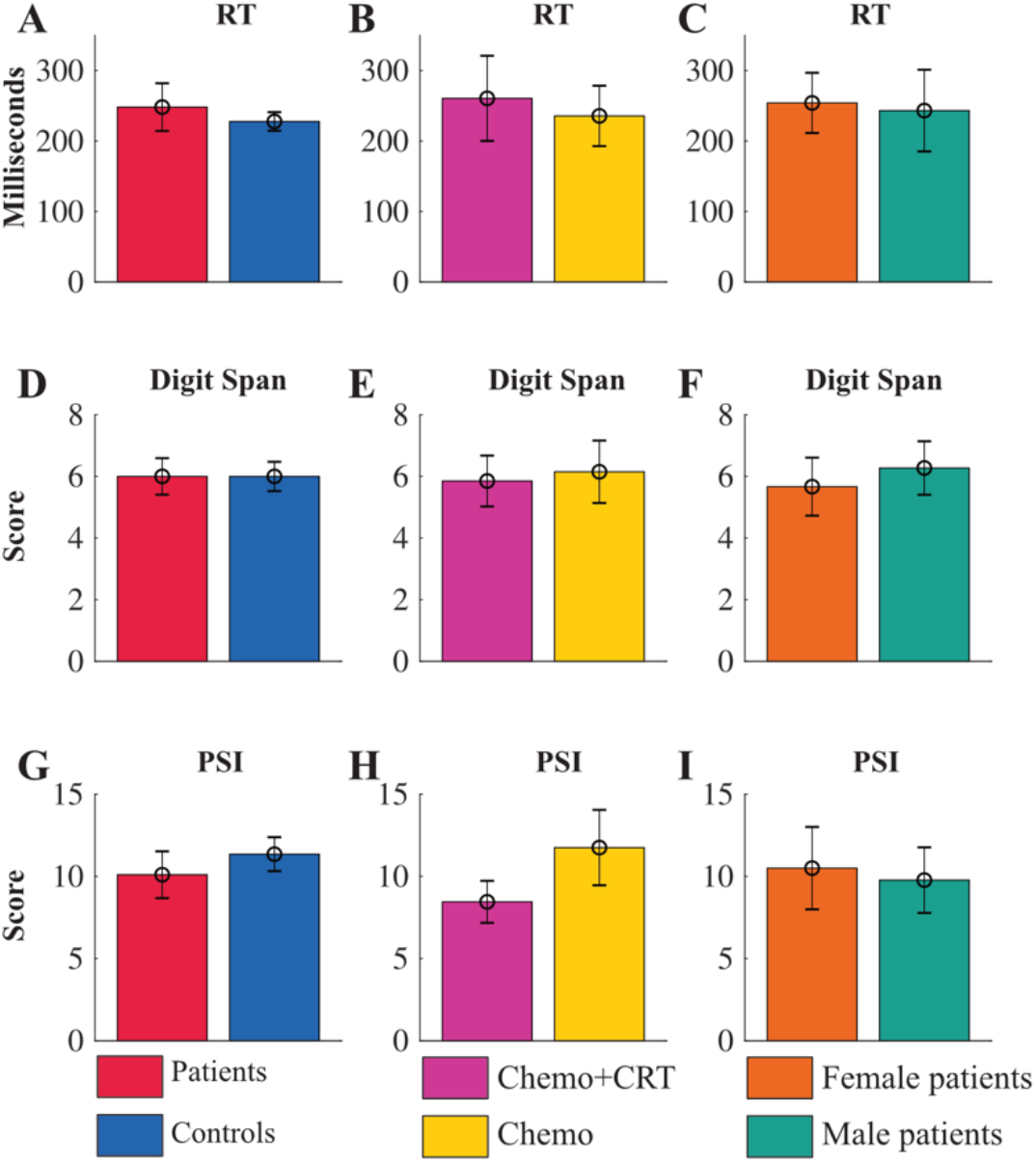
**A-C** show reaction time (RT) for Controls and ALL survivors (A), Chemo+CRT and Chemo (B) and female and male ALL survivors (C). D**-F** show Digit Span for Controls and ALL survivors (D), Chemo+CRT and Chemo (E) and female and male ALL survivors (F). **G-I** show processing speed index (PSI) for Controls and ALL survivors (G), Chemo+CRT and Chemo (H) and female and male ALL survivors (I). Error bars are confidence intervals.

For a measure of proactive and reactive modes of cognitive control, we determined the proactive behavioral index (PBI) from the data of the AX-CPT task. ALL survivors tended to be more reactive i.e., more negative PBI values, and the controls more proactive i.e., more positive PBI values (Figure 4A). The Chemo+CRT group tented to be more reactive compared to the Chemo group (Figure 4B). The difference in PBI between the groups is illustrated by the response time for the AY probe, trials benefitting reactive control, and the BX probe, trials benefitting proactive control (Figure 4D-F). Also, the overall response time in the AX-CPT task was longer for the Chemo+CRT group compared to the Chemo group. The same can be seen in the response time analysis of the CWM-Stroop test where ALL survivors were slower than controls and the Chemo+CRT group responded slower than the Chemo group (Figure 4H). There was also a sex difference in the response time analyses, where female ALL survivors responded slower on both AY and BX trials for the AX-CPT and congruent and incongruent trials in the CWM-Stroop test and with a more reactive strategy (Figure 4C, 4F and 4I).

**Figure 4.**
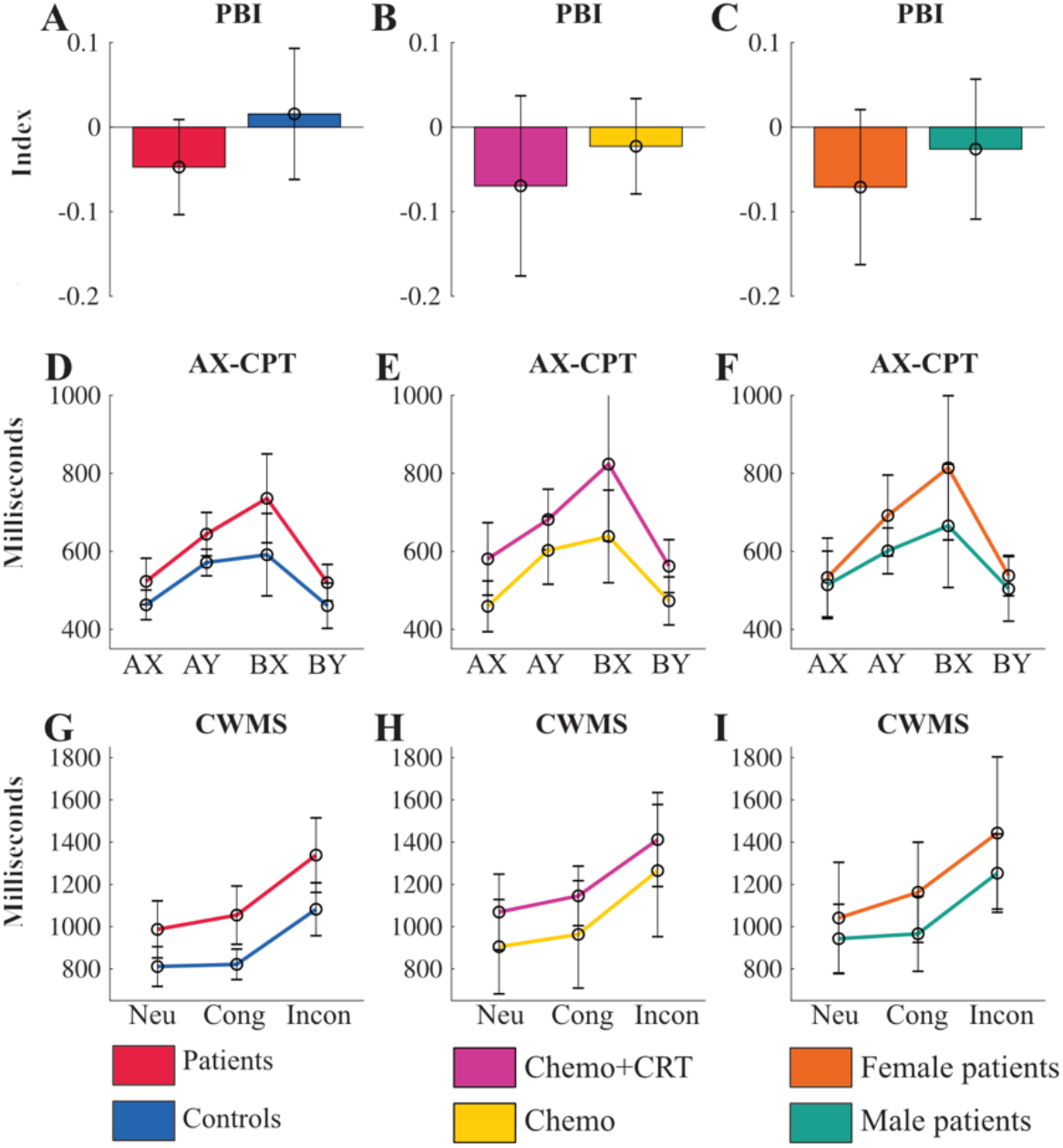
**A-C** show proactive behavioral index (PBI) for Controls and ALL survivors (A), Chemo+CRT and Chemo (B) and female and male ALL survivors (C). D**-F** show response time in milliseconds for the AX-CPT for Controls and ALL survivors (D), Chemo+CRT and Chemo (E) and female and male ALL survivors (F). **G-I** show response time in milliseconds for Color Word Matching Stroop test (CWM-Stroop test) for Controls and ALL survivors (G), Chemo+CRT and Chemo (H) and female and male ALL survivors (I). Error bars are confidence intervals.

### 3.4 fNIRS results

The fNIRS analysis was done on the CWM-Stroop test with exploratory independent t-tests for the mean brain activation in the pooled control conditions and the incongruent conditions for all brain regions of interest. t-values for the difference between ALL survivors vs Controls (Figure 5A-B), Chemo+CRT vs. Chemo (Figure 5C-D), and between female vs. male ALL survivors (Figure 5E-F) are presented. ALL survivors showed a reduced brain activation for the pooled control trails (neutral and congruent) relative to the Controls, especially in the parietal cortex. For the incongruent trials, Controls still activated the parietal cortex more, but patients used the DLPFC cortex more, especially the left aDLPFC. No indication of difference could be detected between the two ALL survivor groups (Figure 5C-D), as well as between Female and Male ALL survivors except for more activity in the right posterior partial cortex for male ALL survivors (Figure 5E-F).

**Figure 5.**
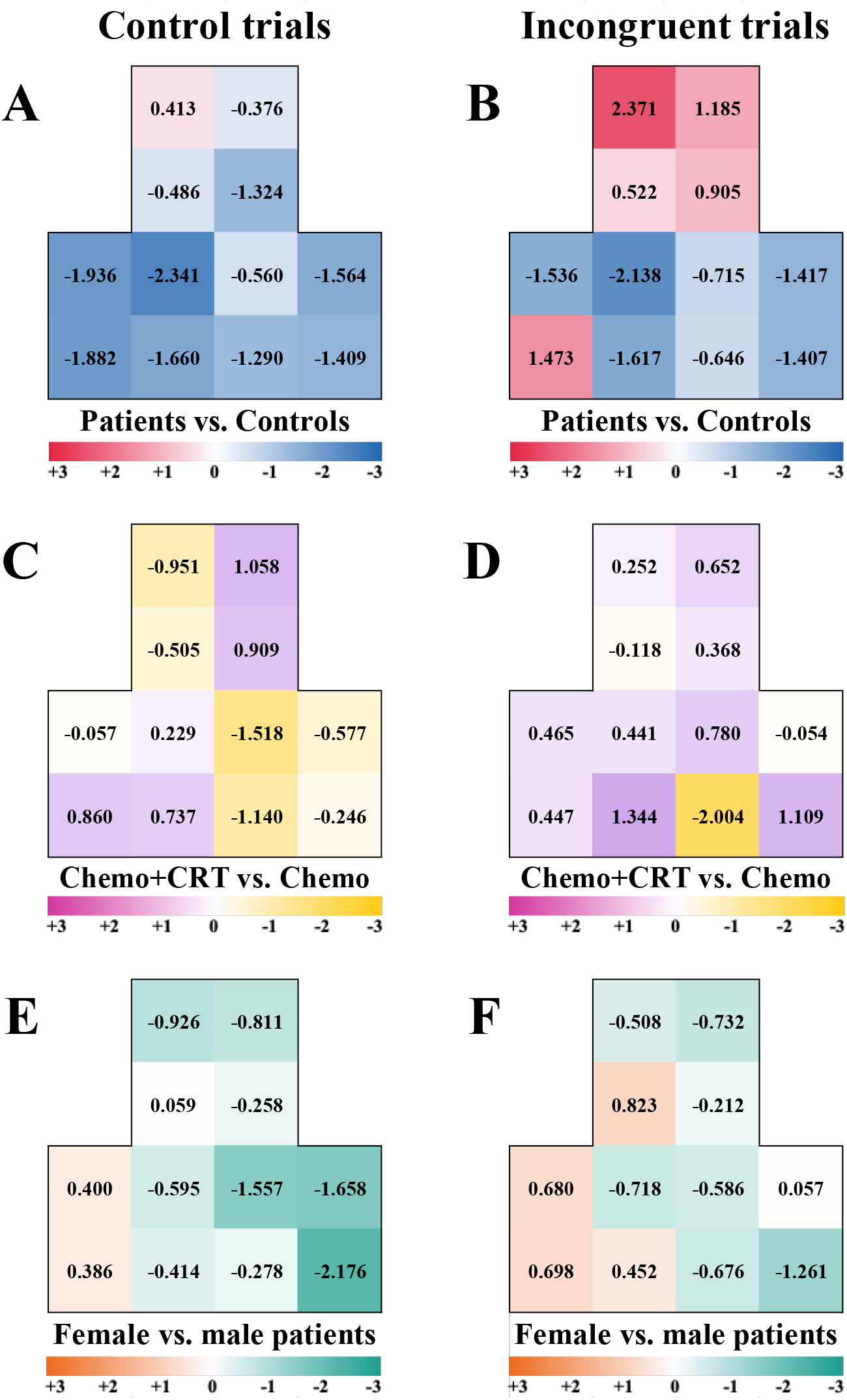
Brain activation differences between groups are presented as t-values Left column mean between Stroop neutral and congruent condition. Right column the incongruent condition. The upper row Patients vs. Controls. Middle columns Chemo vs. Chemo+CRT Lower row Female Patients vs. Male patients. Heatmap is based on independent t-test. Higher values are interpreted as higher oxy-Hb peak for the first group, whereas lower values means higher oxy-Hb peak for the latter group. The heat map reflects t values between –3 to 3. See Supplementary Table 2 for stats.

## Discussion

In this study we used cognitive testing and functional brain imaging to identify and better understand subgroups of adult ALL survivors who suffer from neurocognitive impairments. Overall, on cognitive performance the Chemo group performed similar to the control group, while the Chemo+CRT group performed worse. On mood scales, both ALL survivor groups reported an overall higher level of fatigue and anxiety, but not depression, compared to the control group. During the CWM-Stroop test both ALL survivor groups did not utilize the parietal cortex to the same extent as the control group to solve tasks requiring cognitive control but utilized the DLPFC more during incongruent conditions.

Several reports indicate that adult survivors of childhood ALL experience late complications, including difficulties with fatigue and cognitive functions impacting their ability to work, study, or engage in social activities [10-14, 17]. Additionally, fatigue and anxiety problems have been reported [15, 18, 48].

In this pilot study, although with a relatively small sample size, the heterogeneity of trait fatigue and trait anxiety for ALL survivors, is mostly explained by sex, since most of the female patients report problems (Figure 2F and 2I). The women in both cancer treatment groups also reported a stronger increase in state fatigue during the cognitive test period (2C). A similar sex difference was not detected in the control group (see Supplementary Table 1). An earlier study showed similar sex differences in performance on attention between ALL survivors and controls [11]. In addition, female survivors of childhood ALL reported more pronounced problems with working memory than their male peers [11]. This is in line with the possibility for a hormonal interaction during development with CRT and chemo treatment [11, 17, 20]. However, women have a higher risk of developing problems with fatigue and anxiety than men in the general population as well [49, 50], where these increased risks are also related to sex difference in occupational and social pressure. Since most studies on late effect of ALL survivors focus mainly adolescent survivors and generalized anxiety disorder typically develops in the late twenties for the general population [50], it would be valuable for future studies to investigate the increased risks associated with occupational choices for older ALL survivors.

For the cognitive measures we observed both group (patients vs controls) and sex differences. The Chemo+CRT group exhibited slower performance in both the AX-CPT and the CWM-Stroop tests (Figure 3). Again, women responded slower than men in both patient groups. However, such sex differences were not detected in the control group. The slower response time in AX-CPT and CWM-Stroop test for women in both patient groups should be understood in relation to the similarity in reaction time (Figure 3A-C). Since there was no difference in the pure reaction time test, the difference in the AX-CPT and CWM-Stroop test between the sexes is most probably due to a difference in how information is processed. This is in accordance with reports of individuals with pathological fatigue having slower response time [23, 24].

The PSI from WAIS-IV (Figure 3H) showed the largest difference in processing speed between the Chemo+CRT group and the Chemo group, where Chemo+CRT performed worse, with scores around the 25^th^ percentile for their age. This is in line with previous research on ALL survivors, reporting that individuals treated with CRT as children exhibit slower processing speed [51]. Problems with processing speed have also been related to pathological fatigue [23, 24, 31]. At the cognitive level, the measures of processing speed (Symbol Search and Digit Symbol Coding) are very complex and involve central cognitive domains such as attention, mental and psychomotor operation speed, and visual discrimination [29]. Both attention and visual discrimination are to some degree also involved in the CWM-Stroop test where the Chemo+CRT group also performed worse. Future studies should investigate processing speed by controlling for the cognitive domains mentioned above. This will help determine if the Chemo+CRT group has difficulties with one or multiple cognitive domains and identify aeras where they may need assistance in their daily and professional lives.

The two groups receiving cancer treatment exhibited different patterns of frontal and parietal cortex involvement when solving the CWM-Stroop test. Both groups showed an overall lower functional activity in the parietal cortex during control trials (neutral and congruent) and incongruent trials. Previous studies from our lab have shown that patient groups suffering from pathological fatigue due to traumatic brain injury [23] or exhaustion disorder [24] exhibit less functional activity in the prefrontal cortex during Stroop tasks. However, in this sample the patients utilized the DLPFC more during the incongruent condition. The fact that both ALL survivor groups performed the task similarly but slower and had different functional activity, indicates that they solve the conflict (Stroop effect) differently.

Previous studies have found reduced functional activity for ALL survivors in the parietal and temporal cortex during cognitive tasks compared to controls [20], with an even more reduced activity for female ALL survivors [11]. In this study, no clear sex difference could be determined, although male patients showed slightly more activity in the right posterior parietal cortex. However, this result should be carefully interpreted due to the relatively small sample size (9 women vs. 11 men). In addition, measuring over the posterior parietal cortex may result in nosier data compared to the frontal areas due to its location beneath an area of the scalp with more hair, which can impact the quality of the fNIRS signal.

## Limitations

Since this is a pilot study with a low sample size, no larger generalization should be drawn from the result. Furthermore, the convenience sampling of the controls may not accurately represent the broader population, as participants were chosen based on ease of access rather than random selection. Although we utilized a measure of state fatigue, additional assessments for state anxiety, depression and other factors such as quality of life would have been useful to get an insight into the participants emotional state during the testing day.

## Conclusion

This pilot study aimed to find measurements for evaluating the late complications of childhood ALL survivors on neurocognitive and mood problems in adulthood. The protocol yielded promising results in identifying subgroups that experience neurocognitive and mood problems and we aim to expand upon it in a larger study. As our results indicate increased challenges among female ALL survivors, the interplay between the risk of hormonal interaction with chemotherapy during development and occupational and social pressure during adulthood would be important to investigate in future studies.

## Data Availability

All data produced in the present study are available upon reasonable request to the authors

## Acknowledgements

The authors would like to thank all participants in this study for sharing their experiences and making this research possible.

## Declarations

This study was approved by the Swedish ethics review authority (Etikprövningsmyndigheten – EPN Dnr 2020-01580). All participants received written and oral information about the study, including that their information will be published anonymized as part of a scientific publication, and written consent was obtained prior to the investigation.

No potential conflict of interest was reported by the authors.

## Author contributions

SS: Conception and design. Data collection, Statistical analyses. Interpretation of data. Writing original draft. Review and editing. MJ: Conception and design. Interpretation of data. Writing original draft. Review and editing. GG: Data collection, Interpretation of data, Review and editing. LJ: Interpretation of data, Review and editing. HGK: Conception and design. Interpretation of data. Writing original draft. Review and editing.

