## Supplementary tables and figures for "Using functional near-infrared spectroscopy to explore neurocognitive function in adult survivors of childhood acute lymphoblastic leukemia"

**Supplementary Table 1. Summary statistics**

| | | Woman | | | | Man | | | |
|---|---|---|---|---|---|---|---|---|---|
| | | **Mean** | **SD** | **Min** | **Max** | **Mean** | **SD** | **Min** | **Max** |
| Chemo+CRT | Age | 36.8 | 1.9 | 34 | 39 | 35.8 | 8.6 | 26 | 45 |
| Chemo | | 30.2 | 5.1 | 25 | 37 | 31.7 | 6.3 | 26 | 42 |
| Control | | 35 | 6.1 | 25 | 46 | 33.4 | 7.2 | 22 | 44 |
| Chemo+CRT | Education in years | 14.8 | 1.5 | 12 | 15.5 | 14.2 | 2.7 | 12 | 18 |
| Chemo | | 15.2 | 2 | 13.5 | 18 | 14.3 | 1.6 | 13 | 17 |
| Control | | 15.7 | 2 | 12 | 18 | 15.7 | 2.2 | 13 | 19 |
| Chemo+CRT | VAS pre | 3.6 | 2.1 | 0.8 | 6.2 | 4 | 3 | 1.2 | 8.1 |
| Chemo | | 2.5 | 2.1 | 0 | 5 | 2.8 | 2 | 1.2 | 6.2 |
| Control | | 2.2 | 1.3 | 0 | 4.6 | 3.2 | 2 | 0 | 5.8 |
| Chemo+CRT | VAS post | 6.7 | 2.2 | 3.5 | 8.5 | 4.5 | 3.6 | 1.5 | 9.6 |
| Chemo | | 5.6 | 1.9 | 3.1 | 7.3 | 4.4 | 2.7 | 1.5 | 8.1 |
| Control | | 3 | 1.5 | 0.4 | 6.2 | 3.3 | 2.6 | 0 | 7.7 |
| Chemo+CRT | delta VAS | 2.3 | 1 | 1 | 3.5 | 0.5 | 0.9 | -0.8 | 1.5 |
| Chemo | | 3.1 | 0.7 | 2.3 | 3.8 | 1.5 | 1.8 | -0.8 | 4.6 |
| Control | | 0.7 | 1.5 | -2.7 | 3.5 | 0.6 | 2.9 | -3.5 | 5 |
| Chemo+CRT | MFS | 16.5 | 7.1 | 5 | 24 | 8.6 | 7.7 | 1.5 | 20.5 |
| Chemo | | 16.3 | 6.8 | 9.5 | 23 | 4.9 | 3.1 | 1.5 | 10 |
| Control | | 4.4 | 3.9 | 0 | 9.5 | 5.4 | 4.4 | 0.5 | 13 |
| Chemo+CRT | HAD-A | 7.4 | 2.6 | 4 | 10 | 3.8 | 1.8 | 1 | 6 |
| Chemo | | 10.2 | 2 | 8 | 13 | 4.8 | 2.9 | 1 | 9 |
| Control | | 4 | 3.9 | 1 | 12 | 3.8 | 3.9 | 0 | 13 |
| Chemo+CRT | HAD-D | 3.6 | 1.9 | 1 | 6 | 2.8 | 1.8 | 1 | 5 |

| | | | | | | | | | |
|---|---|---|---|---|---|---|---|---|---|
| Chemo | | 4.7 | 2.1 | 2 | 7 | 1.5 | 0.5 | 1 | 2 |
| Control | | 1.3 | 1.4 | 0 | 4 | 3.7 | 4.1 | 0 | 12 |
| Chemo+CRT | PBI | -0.078 | 0.146 | -0.283 | 0.069 | -0.061 | 0.169 | -0.192 | 0.196 |
| Chemo | | -0.062 | 0.096 | -0.177 | 0.058 | 0.009 | 0.052 | -0.032 | 0.091 |
| Control | | -0.051 | 0.106 | -0.213 | 0.077 | 0.097 | 0.194 | -0.188 | 0.318 |
| Chemo+CRT | PSI | 9 | 1.5 | 7.5 | 11 | 7.9 | 2 | 5.5 | 10.5 |
| Chemo | | 12.3 | 4.1 | 6.5 | 16 | 11.3 | 2.8 | 8.5 | 14.5 |
| Control | | 11.1 | 2.5 | 7 | 15 | 11.6 | 1.8 | 8.5 | 13.5 |
| Chemo+CRT | Reaction Time | 233.3 | 27 | 188.4 | 261.2 | 287.8 | 116 | 193.8 | 482.3 |
| Chemo | | 280 | 75 | 228.2 | 391.2 | 206.1 | 21.8 | 180.5 | 234.5 |
| Control | | 225.5 | 29.8 | 185.6 | 281 | 230.6 | 27.6 | 192 | 290.5 |
| Chemo+CRT | AX | 599.2 | 135.4 | 479.5 | 750.5 | 562 | 137.4 | 391.4 | 762.2 |
| Chemo | | 450 | 71.1 | 373.1 | 527.1 | 466.3 | 112.3 | 391.7 | 664.4 |
| Control | | 491.5 | 80.7 | 393.7 | 659.9 | 427.6 | 70.7 | 349.5 | 526.4 |
| Chemo+CRT | AY | 733.9 | 111.5 | 607.7 | 908.7 | 629.4 | 85.3 | 489.3 | 722.5 |
| Chemo | | 639.2 | 159.5 | 485.8 | 862 | 573.2 | 89.1 | 506.3 | 727 |
| Control | | 605.1 | 68.2 | 508.3 | 707.9 | 530 | 57.6 | 441.9 | 594.4 |
| Chemo+CRT | BX | 884.9 | 276.1 | 619.4 | 1323.7 | 762.7 | 291.9 | 329 | 1065.3 |
| Chemo | | 725.8 | 184.3 | 520.8 | 968.5 | 568.3 | 125.6 | 435.8 | 775.1 |
| Control | | 688.1 | 184.2 | 459.6 | 1006.5 | 473.2 | 223.8 | 242.9 | 838.6 |
| Chemo+CRT | BY | 545.1 | 71.4 | 477.1 | 659.1 | 579 | 120.4 | 462.8 | 777.2 |
| Chemo | | 528.3 | 71.4 | 425.3 | 587.4 | 428.5 | 73.9 | 316.6 | 524.1 |
| Control | | 514.3 | 82.6 | 438.5 | 674.9 | 394.9 | 138.6 | 234.8 | 621.1 |
| Chemo+CRT | Neutral | 1102.7 | 349.5 | 758.9 | 1685.3 | 1035.7 | 127.9 | 817 | 1129.4 |
| Chemo | | 963.8 | 369.3 | 549.9 | 1434.7 | 866.1 | 298 | 538.6 | 1257.3 |
| Control | | 778.3 | 102.1 | 596.8 | 1022.1 | 851.7 | 282.4 | 514.7 | 1315 |
| Chemo+CRT | Congruent | 1178 | 209.4 | 920 | 1374.4 | 1113.5 | 201.8 | 777.2 | 1308.6 |
| Chemo | | 1143.3 | 441 | 528.4 | 1577 | 843.6 | 259.6 | 524.6 | 1178.4 |
| Control | | 822.5 | 117 | 597.5 | 999.6 | 820.3 | 198.4 | 552.8 | 1199.3 |
| Chemo+CRT | Incongruent | 1490.7 | 416.3 | 1021.5 | 1927.7 | 1333.6 | 168.9 | 1108.4 | 1583.8 |
| Chemo | | 1383.8 | 588.1 | 551.8 | 1864.5 | 1186.4 | 343.2 | 672.2 | 1635.2 |
| Control | | 1061.2 | 169.4 | 837.2 | 1316.9 | 1108.5 | 364.7 | 630 | 1657.3 |
| Chemo+CRT | Digit Span | 5.8 | 0.9 | 5 | 7 | 5.9 | 1.5 | 5 | 8.5 |
| Chemo | | 5.5 | 1.7 | 3.5 | 7.5 | 6.6 | 1.2 | 5 | 8.5 |
| Control | | 5.6 | 0.9 | 4.5 | 7 | 6.4 | 1 | 5.5 | 9 |

**Supplementary Table 2. Summary statistics fNIRS t-tests**

| | t-value | df | SD | t-value | df | SD |
|---|---|---|---|---|---|---|
| left DLPFC | -0.486 | 35 | 0.08 | 0.522 | 35 | 0.079 |
| right DLPFC | -1.324 | 34 | 0.065 | 0.905 | 34 | 0.067 |
| left aDLPFC | 0.413 | 33 | 0.078 | 2.371 | 33 | 0.052 |
| right aDLPFC | -0.376 | 35 | 0.074 | 1.185 | 35 | 0.069 |
| left LPPC | -1.882 | 23 | 0.196 | -1.473 | 23 | 0.185 |
| left MPPC | -1.66 | 23 | 0.225 | -1.617 | 23 | 0.256 |
| right MPPC | -1.29 | 25 | 0.239 | -0.646 | 25 | 0.254 |
| right LPPC | -1.409 | 24 | 0.136 | -1.407 | 24 | 0.149 |
| left LPC | -1.936 | 20 | 0.152 | -1.564 | 26 | 0.151 |
| left MPC | -2.341 | 26 | 0.135 | -2.138 | 26 | 0.168 |
| right MPC | -0.56 | 31 | 0.068 | -0.715 | 31 | 0.069 |
| right LPC | -1.536 | 20 | 0.139 | -1.417 | 26 | 0.141 |
| | Chemo+CRT vs. Chemo | | | | | |
| | **t-value** | **df** | **SD** | **t-value** | **df** | **SD** |
| left DLPFC | -0.505 | 16 | 0.077 | -0.118 | 16 | 0.073 |
| right DLPFC | 0.909 | 16 | 0.066 | 0.368 | 16 | 0.084 |
| left aDLPFC | -0.951 | 15 | 0.078 | 0.252 | 15 | 0.059 |
| right aDLPFC | 1.058 | 16 | 0.079 | 0.652 | 16 | 0.057 |
| left LPPC | 0.86 | 12 | 0.055 | 0.447 | 12 | 0.073 |
| left MPPC | 0.737 | 11 | 0.071 | 1.344 | 11 | 0.088 |
| right MPPC | -1.14 | 13 | 0.068 | -2.004 | 13 | 0.081 |
| right LPPC | -0.246 | 10 | 0.06 | 1.109 | 10 | 0.061 |
| left LPC | -0.057 | 11 | 0.087 | 0.465 | 11 | 0.083 |
| left MPC | 0.229 | 13 | 0.083 | 0.441 | 13 | 0.108 |
| right MPC | -1.518 | 14 | 0.06 | 0.78 | 14 | 0.06 |
| right LPC | -0.577 | 12 | 0.082 | -0.054 | 12 | 0.088 |
| | **Female vs. Male patients** | | | | | |
| | **t-value** | **df** | **SD** | **t-value** | **df** | **SD** |
| left DLPFC | 0.059 | 16 | 0.078 | 0.823 | 16 | 0.072 |
| right DLPFC | -0.258 | 16 | 0.067 | -0.212 | 16 | 0.085 |
| left aDLPFC | -0.926 | 15 | 0.078 | -0.508 | 15 | 0.058 |
| right aDLPFC | -0.811 | 16 | 0.08 | -0.732 | 16 | 0.057 |
| left LPPC | 0.386 | 12 | 0.057 | 0.698 | 12 | 0.073 |
| left MPPC | -0.414 | 11 | 0.073 | 0.452 | 11 | 0.094 |
| right MPPC | -0.278 | 13 | 0.071 | -0.676 | 13 | 0.091 |
| right LPPC | -2.176 | 10 | 0.05 | -1.261 | 10 | 0.06 |
| left LPC | 0.4 | 11 | 0.086 | 0.68 | 11 | 0.082 |
| left MPC | -0.595 | 13 | 0.082 | -0.718 | 13 | 0.106 |
| right MPC | -1.557 | 14 | 0.06 | -0.586 | 14 | 0.06 |
| right LPC | -1.658 | 12 | 0.075 | 0.057 | 12 | 0.088 |

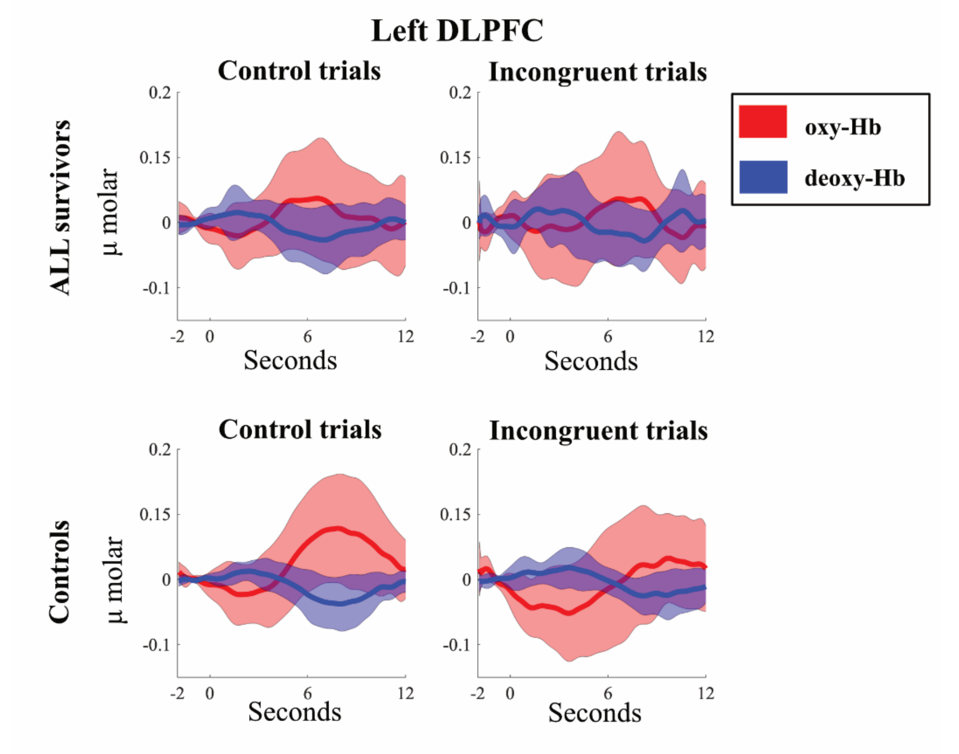


**Supplementary Figure 1. Hemodynamic response CWM-Stroop test.** Mean oxy-Hb (red) and deoxy-Hb (blue) with one standard deviation (light red and light blue) for oxy-Hb and deoxy-Hb respectively for left dorsolateral prefrontal cortex (DLPFC). Control trials involves both neutral and congruent trials. Upper row are ALL survivors and lower row are controls.
